# Spatiotemporal Mapping of Point-of-Care Diagnostic Accessibility: A Data-Driven Pipeline for Point-of-Care Distribution Analysis in Western Uganda

**DOI:** 10.64898/2026.08.28.26361594

**Authors:** Daniel Bergman, Dan Nyehangane, Lonni Besançon, Maria Podkorytova, Vasiliki Tsoumari, Dimitrios Staikoglou, Angella Kimuli Namyalo, Muganzi Richard Rukakaza, Patrick Ogwok, Claire Nankoma, Tobias Alvén, Juliet Mwanga-Amumpaire, Giulia Gaudenzi

## Abstract

**Background:** Accurate diagnosis is foundational to effective clinical decision-making and patient outcomes. Availability to essential *in vitro* diagnostics within the primary health care in Uganda was 10-15% in 2007. Tools to analyse and visualise diagnostic data are essential across clinical, commercial, and policy contexts. Here, we present a novel visualization pipeline using diagnostic availability data from Western Uganda as a case study, alongside a generalizable, data-driven framework for POCT distribution analysis.

**Method:** All primary healthcare centers owned by the Ugandan government in the Western Region of Uganda were submitted to a questionnaire concerning current availability of POCT from the Essential diagnostic List 2 part 1a and 1b, and the African laboratory inventory done by African Society of Laboratory Medicine and AfricaCDC. The data from the questionnaire was then linked to open source geodata provided by TomTom, and population data to calculate and visualize the accessibility of captured POCT.

**Findings:** Availability of POCT Malaria is almost 100%, HIV 68-90%, and >30% for a majority of the POCT in the EDL-2 panel. 90% of the population in Western Region live within 1 hour by car from most of the essential POCT. Figures in the complementary web-based application visualize the accessibility of POCT for Western Uganda. Diagnostic deserts are visualized.

**Interpretation:** Access to POCT at primary health care facilities in western Uganda has expanded substantially over the past decades. The geo-mapping tool presented here could inform policy decisions on strengthening diagnostic capacity at the national, regional, and provincial level.

**Funding:** Swedish Research Council and Infravis

All supplementary materials and a preprint of this submission are available on our OSF repository https://osf.io/j7puk/.

## Introduction

Accurate diagnostics are the cornerstone of modern medicine, essential for precise assessments and effective patient care. Over the past decade, the World Health Organization (WHO) has increasingly emphasized their critical role, notably through the establishment of the “List of Essential In Vitro Diagnostics” (EDL), a framework designed to enhance diagnostic accessibility and prioritization worldwide. This commitment was further reinforced by World Health Assembly (WHA) Resolution 76.5, which calls for strengthening diagnostic capacity and urges Member States to integrate national diagnostics strategies into their healthcare plans. From a global health perspective, point of care tests (POCT) are, in many settings, the only available diagnostic options. Accessible and reliable POCT enables earlier disease detection, timely initiation of treatment. In accordance with the above, the African Society for Laboratory Medicine (ASLM) has together with AfricaCDC started an initiative to map laboratory capacity in Africa.

Uganda is a low-income country where previous mapping of diagnostic tests has indicated low availability, specifically in primary health care settings.^1–4^ Shortage of diagnostics tests have been reported in other Sub-Saharan African countries.^5,6^

This study gathered data on the availability and accessibility of POCT in Ugandan primary health care settings. In addition to this gathered data, we further developed a spatiotemporal visualization and analysis pipeline. This data-driven software provides several benefits to our research objectives of mapping diagnostic capacity of Western Uganda, but is also likely to advance the planning of developments to come for the region as well as provide the overall global-health community, from researchers to policy-makers, with an easy, interactive, and visual exploration of the needs of different regions for POC diagnostics.

## Method

### Study design

This is a descriptive study on availability of POC diagnostics at primary health care centers (HC) in Western Region of Uganda.^7^ The study has a temporal component of both investigating which tests that were available on the day the questionnaire was filled out and which tests that were available on the 31st of December 2019. This date was chosen because it is just prior to the Covid-19 pandemic, with the purpose of investigating differences in diagnostic capacity pre and post Covid-19. Data was collected via a REDCap questionnaire (available on our OSF.io repository) between 25th of November 2024 and 18th of March 2025.

The Western Region of Uganda is one of four regions with a population of 11,5 million inhabitants. Primary health care in Uganda is organised into basic primary health care (HC II) and advanced primary health care (HC III and IV). ^7^

#### Data collection

All data were collected using a standardised questionnaire filled out through an interview with a focal person at each HC. This focal person being laboratory personnel at HC’s with a laboratory or nurses on HC without a laboratory (using only POCT). Interviews were conducted either in person or through telephone. No instruments or equipment has been observed by the surveyor.

The questionnaire captured data on the availability of tests from EDL-2 1a and 1b and supplemented by a list of tests requested by the African CDC (Center for Disease Control and prevention) and ASLM (African Society for Laboratory Medicine).^8^ In addition to questions on available tests on the day the questionnaire was filled out and 31st of December 2019, information on quality control and how the HC handled diagnostic tests they could not perform was collected. Data entry and management was done by a data entry team in Mbarara. Data was entered to the REDCap (Research Electronic Data Capture) App for Android and then uploaded to the REDCap database when internet connection was available. Statistical analysis was performed using RStudio (version 2025.09.0+387) and Google Sheets.

#### Inclusion criterias

HC II-IV in the Western Region of Uganda owned by the Ugandan Government.

#### Exclusion criterias

Health facilities not characterised as HC II-IV, like hospitals and specialised clinics. Health facilities with ownership other than the Ugandan Government. Health facilities outside of the Western Region of Uganda.

#### Geographical data

Data on geographical coordinates of each HC was primarily obtained using a free web based tool https://www.gpsvisualizer.com/, that scans google maps for the coordinates of requested locations. These coordinates were then reviewed to assess if they deemed accurate. 664 HC coordinates were obtained automatically. 245 coordinates of HC were obtained by manually searching google.com/maps and mapacarta.com. The coordinates of 23 HC were not obtained.

#### Data-Driven Visualization Pipeline

Collected data was cleaned and filtered by the HCs which were inside the borders of Western Uganda. Facilities were extracted for each of the chosen POCT (see the list of POCT in Table 1). For each of the possible POCT we built service areas based on network analysis. The analysis was performed in ArcGIS Pro software, version 3.6.3, based on TomTom road network data and using Dijksta’s algorithm. The result of the analysis was integrated with the open-source population data. “2.5D maps” was created for the relatively available POCT. These maps consist in using color to map travel time to a specific HC for the POCT of interest (e.g., in Figure 2 and 3) and using the height of the unit to depict population density. The map can efficiently convey both a variable of interest and its importance when considering population density, allowing both a quick overview of the data and a more detailed analysis (e.g., it is easy to see if there exist highly-populated places with difficult access to diagnostics).^9^ Two interactive graphs were created, a scatterplot with the overall availability statistics on a health center level for the period before and after the Covid-19 pandemic and a radar graph which illustrates the statistics derived from the accessibility maps.

**Table 1.** Number of people living within distance in “minutes by car” from each test. hCG = Human Chorionic Gonotropin, Hb = Hemoglobin, Ag = Antigen.

| Minutes by car to |  |  |  |  |  |  |
| --- | --- | --- | --- | --- | --- | --- |
| POC-test | 0 - 10 | 10-30 | 30 - 60 | 60 - 90 | 90 - 120 | more than 120 |
| Albumin in urine | 656 329 | 2 794 879 | 3 439 038 | 1 732 948 | 1 088 602 | 1 012 917 |
| Syphilis antibody | 3 872 432 | 5 443 868 | 1 181 125 | 97 559 | 49 030 | 80 698 |
| HIV & Syphilis | 5 125 188 | 4 859 766 | 644 344 | 41 098 | 16 529 | 37 787 |
| HIV CD4 | 3 223 044 | 5 569 696 | 1 604 192 | 188 207 | 58 532 | 81 042 |
| Hepatitis B | 3 951 243 | 5 423 898 | 1 191 422 | 94 860 | 19 788 | 43 501 |
| HIV1 and HIV2 | 5 085 360 | 4 896 053 | 648 208 | 40 149 | 16 535 | 38 409 |
| Malaria | 5 608 234 | 4 572 032 | 451 356 | 38 119 | 16 535 | 38 437 |
| Urine Dipstick | 3 664 760 | 5 511 167 | 1 303 842 | 115 184 | 49 060 | 80 701 |
| Glucose in urine | 3 659 193 | 5 516 763 | 1 303 842 | 115 184 | 49 060 | 80 671 |
| Ketones in urine | 3 659 193 | 5 516 763 | 1 303 842 | 115 184 | 49 060 | 80 671 |
| Glucose in blood | 3 668 503 | 5 481 477 | 1 335 062 | 121 535 | 49 613 | 68 523 |
| hCG | 4 372 168 | 5 196 400 | 974 289 | 71 570 | 58 090 | 52 196 |
| Bilirubin in urine | 3 026 867 | 5 374 188 | 1 908 756 | 269 114 | 65 248 | 80 540 |
| Cryptococcal Ag | 3 264 015 | 5 308 669 | 1 763 341 | 247 161 | 56 289 | 85 238 |
| Blood typing | 3 068 726 | 5 556 456 | 1 674 758 | 268 657 | 73 250 | 82 866 |
| Hb in Blood | 3 187 982 | 5 647 677 | 1 607 540 | 150 838 | 50 069 | 80 606 |
| Hb in Urine | 3 611 460 | 5 523 110 | 1 343 822 | 116 616 | 49 057 | 80 647 |

The results of the analysis were integrated in the interactive GIS app. The app was created using ArcGIS Online and ArcGIS Experience Builder. Previous studies typically follow a single analytical pathway, assuming one accessibility problem and identifying an optimal solution.^10–12^ Those approaches are limited under uncertainty and are not well suited to unanticipated disease outbreaks. Instead, we retain a diverse set of tests and enable flexible interaction with the data. This allows the analysis to be rapidly adjusted and supports responses to questions that cannot be specified in advance.

The application includes eighteen service area maps and forty two types of care diagnostics, combined through filters for two time periods. Moreover, the 2.5D maps combine accessibility, defined through service areas, with population data to provide clear and engaging visual communication. Accessibility maps for 17 priority POCT were created and integrated through a vector overlay in ArcGIS Pro. Areas where accessibility exceeded 1 hour for at least one of the 17 POCT was identified. These areas were intersected across all POCT and mean travel time was calculated for the full set (Figure 4), to construct a map of diagnostic deserts. Limited accessibility for a single disease produces a relatively low mean value, whereas limited accessibility across multiple POCT indicates a more systemic problem.

### Ethical concern

No personal, biological or sensitive data has been gathered during this research project. Ethical approval from Mbarara University of Science and Technology was obtained with administrative no: MUST-2023-1161 and Uganda National Council for Science and Technology with reg no: HS3718ES.

## Results

A total number of 932 HC were surveyed, of which 477 were HC II, 387 HC III and 68 HC IV. 37·3% (348) of all HC reported that they have a clinical laboratory performing tests other than POCT (microscopy for malaria included in POCT). 20·5% (98) were at the level of HC II, 55·5% (215) of HC III and, 51·4% (35) of HC IV. Geographical coordinates were deemed accurate for 886 HC. POCT were widely available, primarily Malaria and HIV-diagnostics (Figure 1). In HC II and III less than 1% reported that a specific test is available but temporarily out of stock. 52·2% of HC IV reported stock outs of Covid-19 tests. 4·3% and 2·9% of HC IV reported stock outs of Hemoglobin in blood respectively combined HIV/Syphilis test. Apart from the above mentioned, no other “stock outs” were reported.

**Figure 1.**
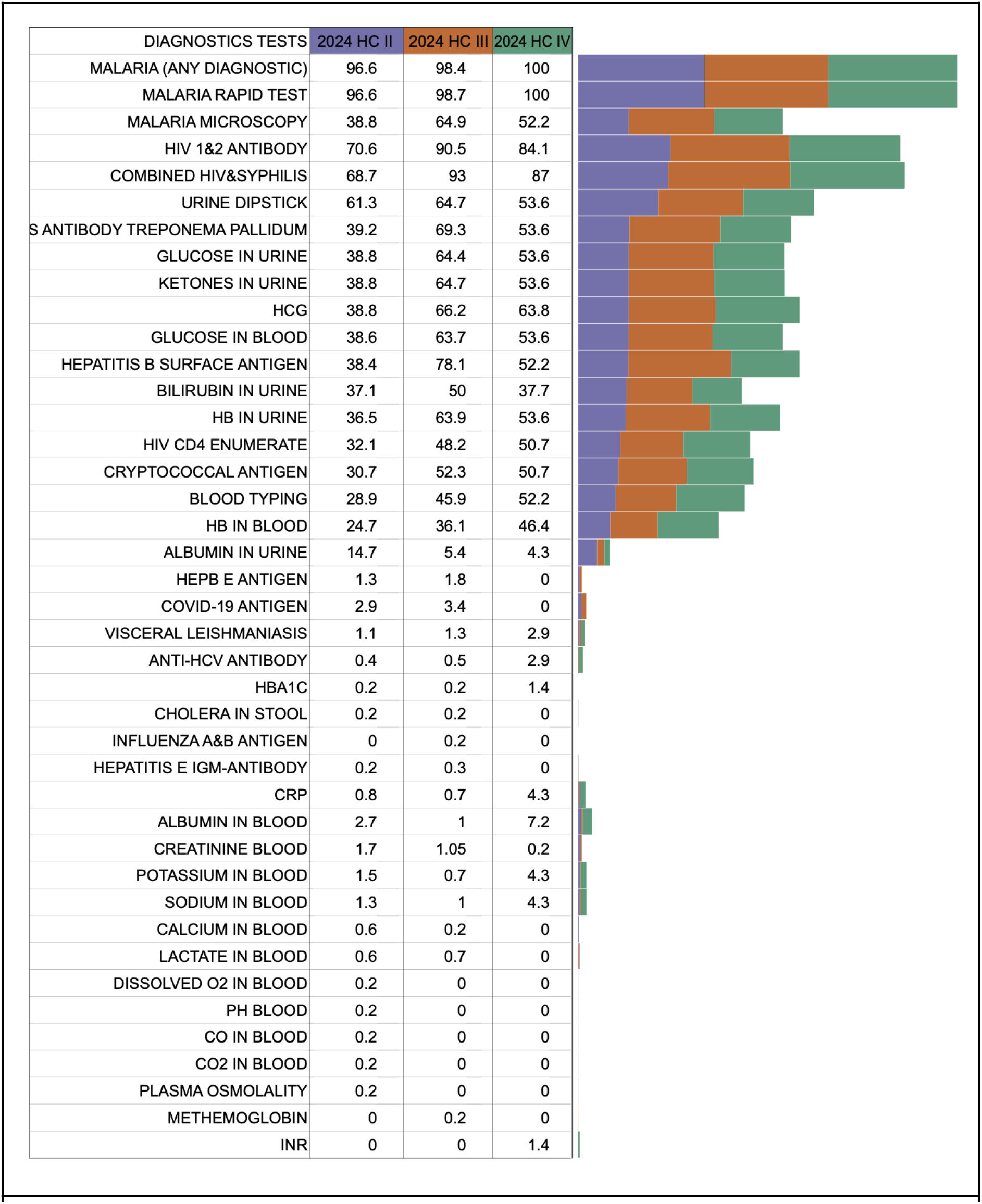
Percentage of health centres by tier with each POCT available at the time of the study.

‘2.5D maps’ (Figure 2 and 3) present distance in minutes by car to a specific test and population density. The distance in minutes takes into account the quality of the roads and includes secondary rural roads in the analysis, so the analysis rather overestimates than underestimates the availability of tests, especially in connection to seasonal change in quality of the roads. Figure 2 and 3 show that the southwest of the Western Region is fairly densely populated and that driving time to a malaria-test is in general <10 minutes, but for HIV CD4-testing the general time is 10-60 minutes. Densely populated areas have better access to diagnostic services. Accessibility maps for all existing POC-tests, will be found in our online extension of this printed article (https://arcg.is/1W4Sva).

**Figure 2.**
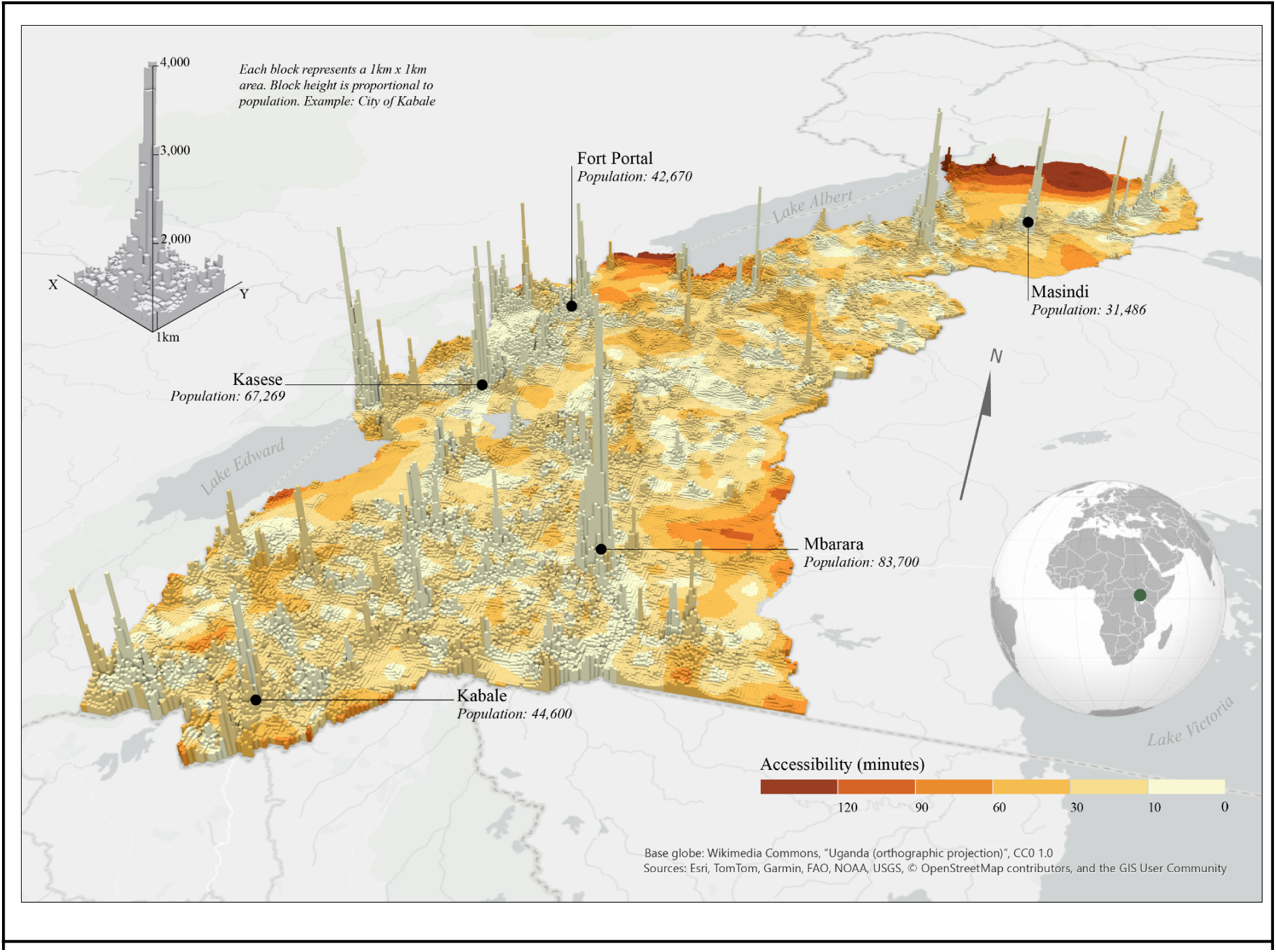
Population and time by car to the closest HC with HIV CD4 POCT.

**Figure 3.**
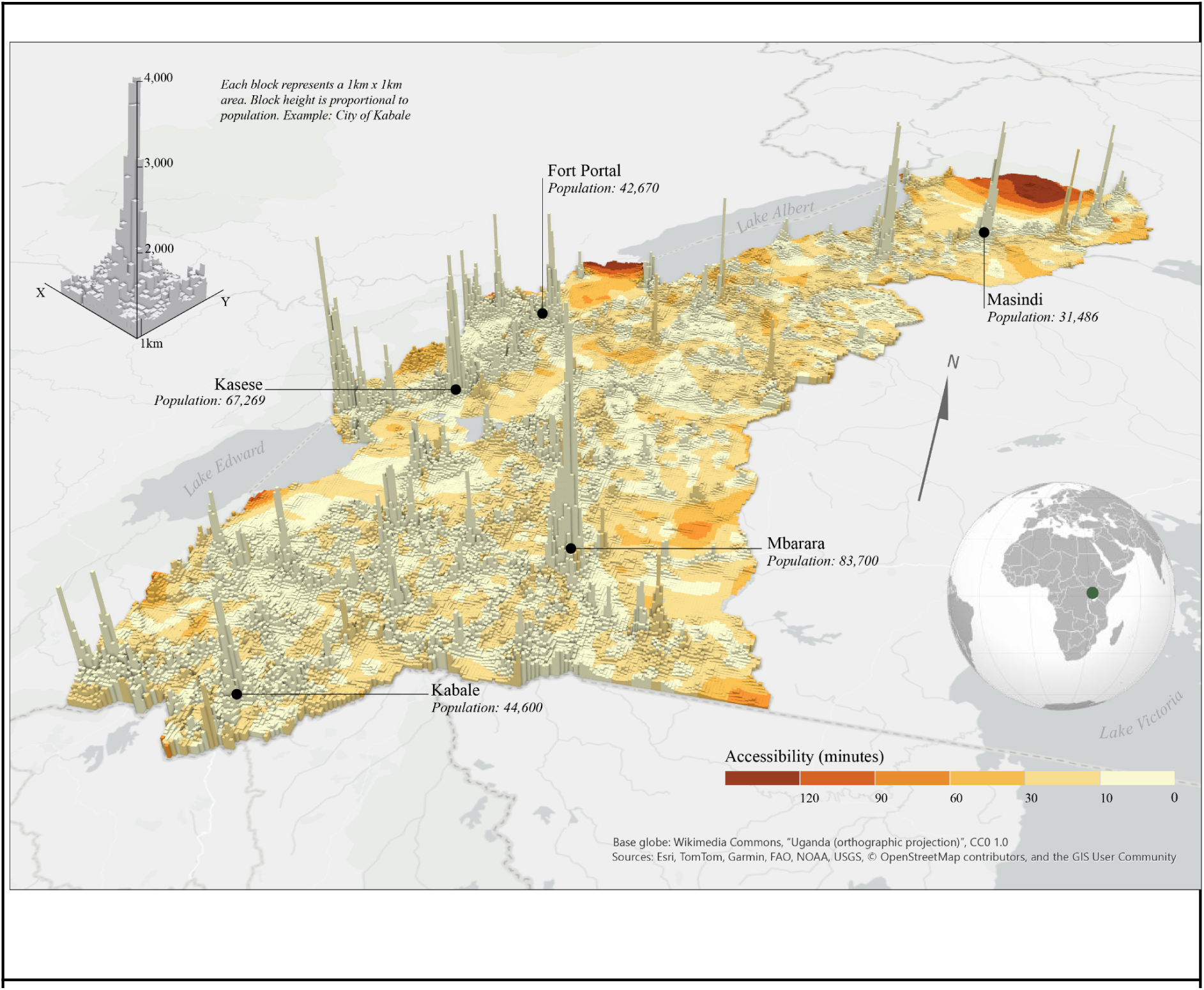
Population and time by car to the closest HC with Malaria POC-testing.

Approximately 6% of the population of Western Region of Uganda lives more than 30 minutes by car from a HC with Malaria-testing and 7% more than 30 minutes by car from a HC with HIV-testing. Many of the tests on the EDL lists are reachable within 60 minutes for a clear majority of the population.

The map of diagnostic desserts (figure 4) underscores the spatial inequality in POCT in the Western Region of Uganda. The largest areas with low availability of diagnostic services are located at the periphery of the region, particularly in the sparsely populated north. The most prominent area in the north corresponds to national parks and wildlife reserves, where low population density and regulatory constraints limit infrastructure development. Additional areas of low availability are observed along regional borders, suggesting potential issues of transport accessibility that require further investigation. These patterns differ by border type. Low availability at national borders may indicate a substantive access constraint, whereas similar patterns at intrastate borders may be less critical, as populations can access services in neighbouring administrative units within the same country. In the latter case, the pattern reflects the modifiable areal unit problem (MAUP), indicating the need for more detailed data to support interpretation.

**Figure 4.**
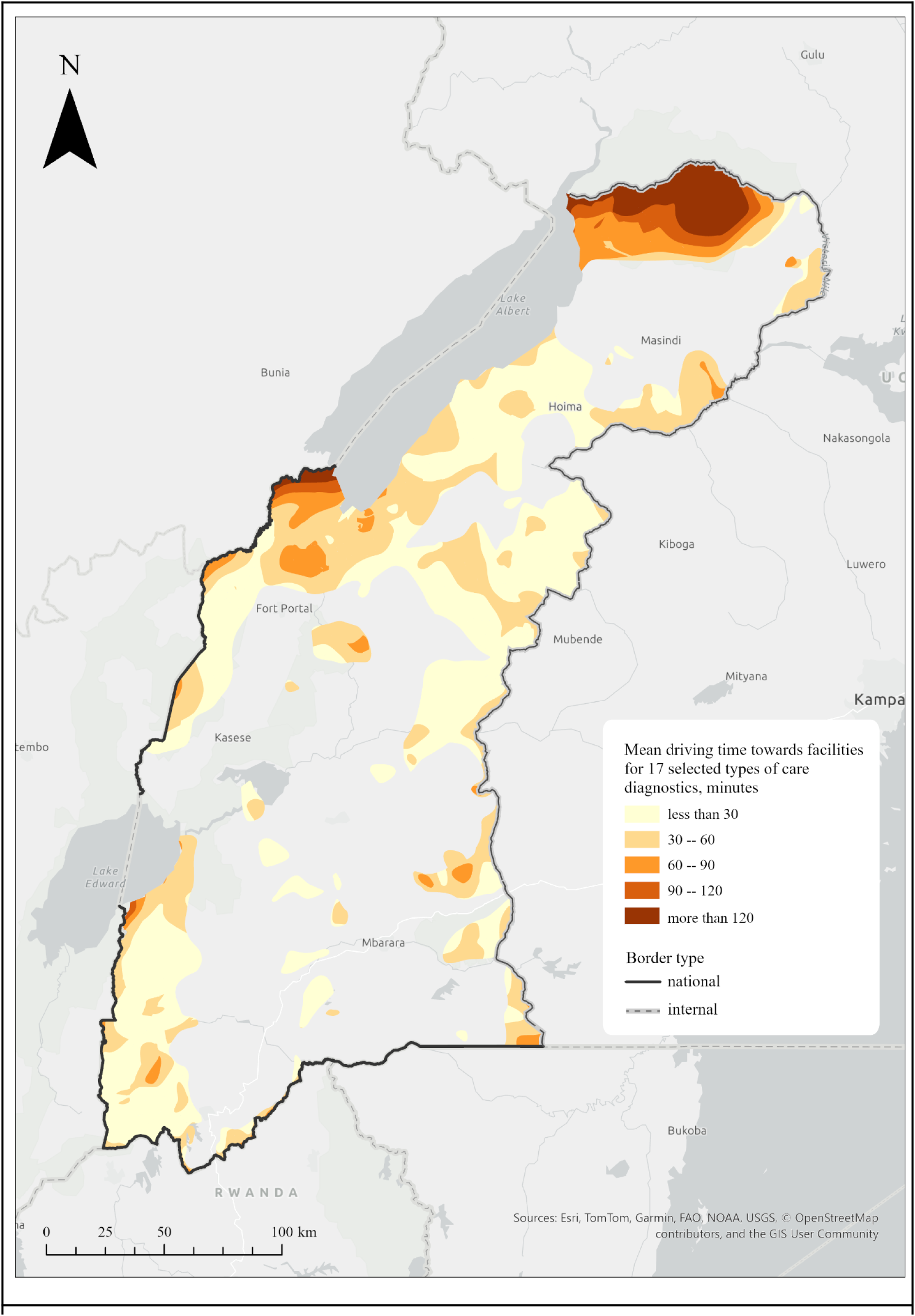
Mean driving time to 17 commonly existing EDL-POCT, where at least one POCT has a driving distance >60 minutes. In grey areas all 17 POCT are found within 60 min driving distance.

In the total set of HC 48·9% (456) sent samples to a laboratory, to perform a test that the HC could not themselves perform. For each health tier, HCII 41·7%, HC III 66·1% and, HC IV 54·4% respectively sent samples to a central or other laboratory. The median and mean number of sent sample per week was, 20 (IQR 20) and 24·5 (SD 29·9) for HCII, 15 (IQR 10) and 17·4 (SD 10·5) for HCIII, and 25 (IQR 10) and 26·1 (SD 10·9) for HCIV. The median turn around time (TAT) being 10 (IQR HCII 7, HCIII 7 and HCIV 4) days for all three health tiers and the mean TAT being 10·3 (SD 4·4), 10·5 (SD 3·0) and 12·9 (SD 4·8) days for HC II, HC III and HC IV respectively. 49·7% (463) of all HC referred the patient to a different clinic if they could not perform a diagnostic test that they deemed necessary, instead of sending a sample to a central laboratory.

Data pre- and post-pandemic availability of POCT demonstrate a general increased availability of POC-diagnostic (Figure 5). The difference between pre- and post-pandemic is relatively small though.

**Figure 5.**
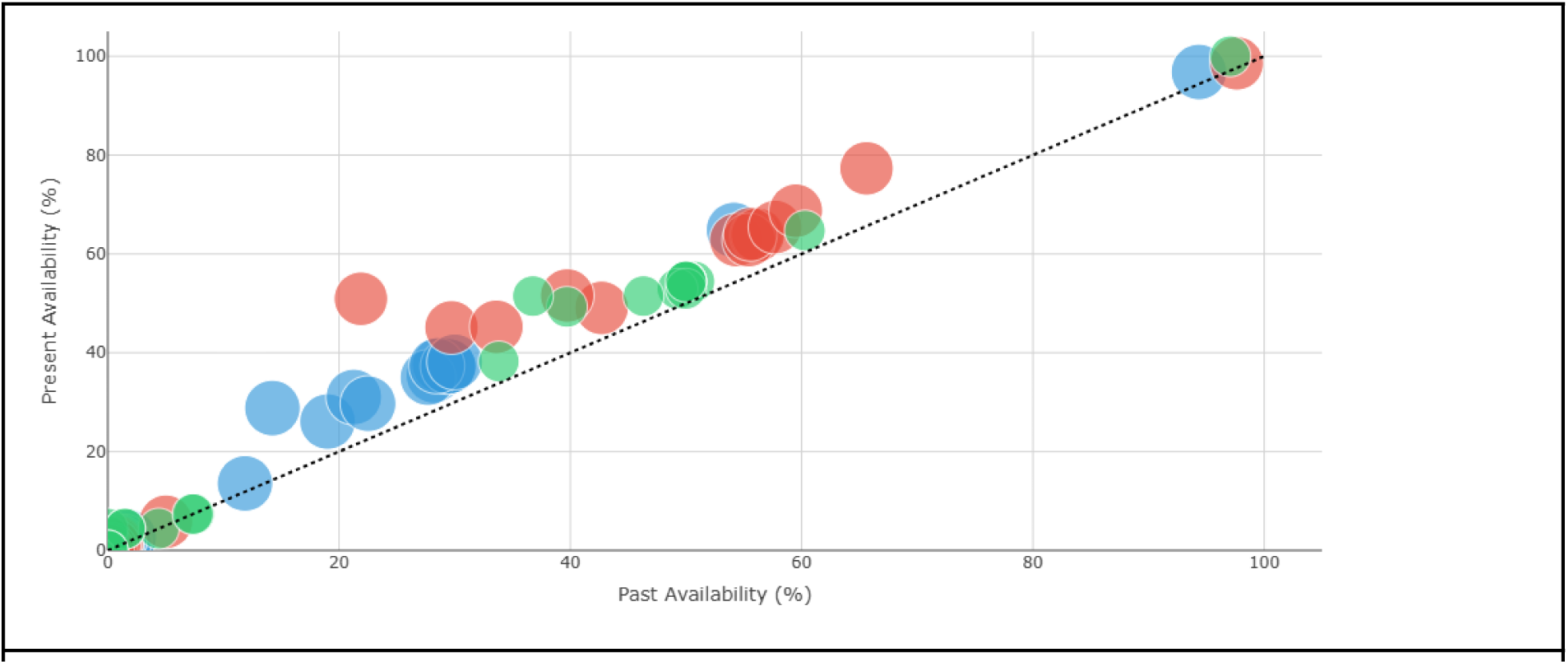
Availability of EDL POC-diagnostic test in HC II-IV, pre- and post Covid-19 pandemic. Blue = percentage of specific test available at HC II, Red= percentage of specific test available at HC III, Green = percentage of specific test available at HC IV. Bubble sizes proportional to the number of health centers. Find interactive diagram at https://arcg.is/1W4Sva

**Table 2.** HC reported that they perform any kind of quality control on their diagnostic tests.

|  | Reported any quality control | Known pos/neg | Specific test |
| --- | --- | --- | --- |
| All HC | 32.9% (307) | 27.7% (242) | 8.9% (83) |
| HC II | 32.9% (157) | 22.0% (105) | 13.0% (62) |
| HC III | 29.4% (114) | 26.4% (102) | 3.9% (15) |
| HC IV | 52.9% (36) | 51.5% (35) | 8.8% (6) |

The most common control reported was UVRI EQA (Uganda Virus Research Institute External Quality Assurance), reported by 77 of 83 HC using a control. 64 HC had laboratories ranked according to the ASML SLIPTA program, 38 HC with 0 stars, 13 HC with 1 star, 7 HC with 2 stars, 3 HC with 3 stars and 3 HC with 4 stars.

## Discussion

Availability of diagnostics tests in general has improved over the last 20 years, with dramatic improvements of Malaria and HIV diagnostic tests. 2007 around 15% of all HCII had diagnostic tests for malaria and 5% had HIV-tests, in 2024-25 96% had malaria tests and close to 69% had HIV-tests.^1,13^ At all levels of primary health care the availability of in vitro diagnostic tests has improved compared to data from 2007. Availability of diagnostic tests is also better compared with data from Kampala in 2011 and similar to relatively recent comparable data in Kenya.^2,14^ The arsenal of POCT available in primary health care has expanded impressively over the past 20 years, with a continuing positive trend since the Covid-19 pandemic. It is not possible from this dataset to say that the pandemic had a positive effect on the availability of POC-diagnostics, rather that it fits the trend over the last 20 years. It should be noted that the improved availability of diagnostics tests in this study solely is based on POCT, tests sent to a clinical laboratory are not included in this improved statistics.

Availability of diagnostic tests at each HC does not necessarily correspond to how accessible those tests are to the population of the Western Region. Previous studies have analysed accessibility to care or diagnostics in different ways. In this study we chose to analyse this through distance in minutes by car. Studies from Ghana have conducted similar accessibility analysis and used the speed of a motorized tricycle 20 km/hour, a study carried out in Madagascar, based travel time on walking.^10,15,16^ By incorporating population distribution as a variable (figure 2, 3 and https://arcg.is/1W4Sva), the analysis moves beyond simple facility mapping to reveal where diagnostic capacity is, and critically, where it is not, aligned with population need.^9^ For policy makers or other administrative units this could be used to analyse if there are any diagnostic underserved areas and where they exist. Accessibility maps coupled with the interactive map provides a clear overview of HC that are located in a specific region of interest and which POCT they provide.

The COVID-19 pandemic increased uncertainty in cross-border interactions, reducing the transparency of national borders and disrupting everyday practices in local communities. As a result, diagnostic care deserts located along national borders, such as the border with the Democratic Republic of the Congo, may become spaces of exclusion. For example, the Nyakasenyi area, situated between the national border and a nature reserve, has experienced heightened stress as reduced border transparency has constrained mobility.^17^

During the pandemic, the opportunity of crossing national borders became uncertain, whereas internal administrative boundaries remained relatively open. This distinction underscores the importance of identifying diagnostics deserts at national borders, where restricted mobility can intensify barriers to accessing healthcare. Limited accessibility to POCT is also observed in relatively populated areas. For example, the area south of the Kisoro–Kabale road, in the Southern part of the Western region of Uganda, has a mean travel time of approximately 45 minutes for key diagnostic services. Even a single additional low-level healthcare centre located closer to this area could substantially improve access.

The map of POCT deserts reveals a heterogeneous pattern of diagnostic availability. Spatial analysis helps to identify areas where targeted interventions may be feasible; however, such actions should be validated with local expertise and considered in relation to existing mobility practices. The data driven tool presented in this study could be used to locally plan diagnostic capacity.

This study tries to capture accessibility and population density in a static visualisations and couple them with interactive maps to enable researchers, healthcare specialists and urban planners to identify the most vulnerable areas from the point of availability of care diagnostics and most underrepresented types of care diagnostics. Similar previous accessibility studies are not interactive and leave out population density from the visualization.^10,15,16,18^ However, parts of our current pipeline with those tools relies on closed-source projects that limit the applicability of our pipeline to researchers and stakeholders with available licences of the software we use. Having identified through this project that the resulting data-driven analysis pipeline is of particular use to researchers and stakeholders in global health. The main limitation of the approach is that it uses commercial software, so it can’t be reproduced outside a licensed environment. The further development of the project should focus on moving towards open source and open data solutions, especially considering the limited resources of the field. We thus plan to now work on an Open Source version of our analysis that would then be available to all researchers working with similar data. A key limitation is the heterogeneity of digital geographical data and limited access to some types of it. Many fields require open, standardised data on human mobility to move beyond hypothetical routes derived from road networks and instead capture actual movement patterns.^19^

Not all POCT are needed at every HC. Ultimately, health care facilities must have the capacity to deliver appropriate management for the results of diagnostic tests that they use, whether this entails initiating treatment or referring the patient to a higher health tier. Of the HC II almost 60% referred patients to a different health facility in cases where they assessed that they needed laboratory tests not available at their facility, which could imply an understanding of the kind of medical problems the HC could manage even with a test-result.

Uganda has since 2011 developed and implemented a system of laboratory hubs, to serve surrounding HC with laboratory services.^20^ Approximately half of the HC in our study sent tests to a central laboratory. For tests that were sent the TAT was approximately 10 days, indicating that no test done with an expedient need could be sent. POCT are in many cases the only feasible solution, especially for time-critical diagnostic tests, particularly in rural settings. Malaria stands for almost 9% of all deaths in Uganda and 10% of all disability adjusted life years (DALY), a diagnosis needing expedient management.^21^ POCT have made it possible to manage a good portion of all malaria cases at primary health care level.^22^

From the self reported data in this study quality control of the in vitro tests at the HC mounts up to one third in the whole sample and 52.9% in HC IV. Considering this, the quality of POCT used in most HC could be questioned. This result indicates scope for improvement and further studies on the need for better quality control.

A methodological limitation of this study concerns the ascertainment of geographical coordinates. Rather than recording coordinates prospectively in the field, the authors derived them retrospectively from openly available online mapping tools (Google Maps; Mapacarta). This approach introduces potential spatial imprecision and may compromise the reproducibility and reliability of any location-dependent analyses. Sites were verified at district level, with a subset additionally confirmed at sub-county level; however, some residual coordinate error is probable. This trade-off was accepted in order to maximise sample size and operational feasibility, acknowledging that a smaller, more precisely geo-located sample would have been the alternative.

Potential misreporting by survey respondents is an additional source of uncertainty. The survey was conducted via telephone and POCT were therefore not directly inspected. The model has inherent geographical limitations at administrative boundaries. Districts bordering other regions of Uganda are likely to carry elevated error values, as health facilities located immediately across regional boundaries were not captured in the sampling frame. A similar limitation applies to populations residing in proximity to hospitals or clinics operated by non-governmental organisations, which were likewise excluded from the study. Taken together, these boundary effects mean that the true accessibility of diagnostic services is likely to exceed the estimates reported here. Simultaneously, travel time analysis in this study may overestimate accessibility. Effectively these limitations balance each other.

## Conclusion

The availability of POC-diagnostics has increased substantially over the past 20 years.^1^ Very few people in the Western Region of Uganda live more than an hour (with car) away from a HC with a possibility to test for malaria, and more than 50% of the population live less than 10 minutes (with car) away from a HC with the possibility to test for malaria. Many of the tests on the EDL-diagnostic list for POCT are relatively available in Western Uganda. Our method of analysing and presenting the availability and accessibility of diagnostic tests could be a basis in policy management on diagnostic expansion. Even though diagnostic tests are essential to modern medicine, the personnel reading the results must have some means to manage the patient. Which tests that then should be available at different health tiers is a discussion beyond the scope of this article. With this in mind data from this study can be used to plan future expansion of diagnostic capacity and to ensure its availability to the entire population, thereby increasing opportunities for appropriate treatment. An interesting development of the data driven pipeline presented in this paper is to develop it into a live dashboard to monitor low supplies or stock outs in real time.

https://www.elsevier.com/researcher/author/policies-and-guidelines/credit-author-statement

## Contributor Roles Taxonomy (CRediT)

**Daniel Bergman:** Conceptualization, Methodology, Formal analysis, Data Curation, Writing - Original Draft **Dan Nyehangane:** Conceptualization, Investigation, Resources, Writing - Review & Editing, Project administration **Lonni Besançon:** Visualization, Writing - Original Draft, Methodology, Resources **Maria Podkorytova:** Visualization, Writing - Original Draft, Methodology **Vasiliki Tsoumari:** Visualization, Methodology **Dimitrios Staikoglou:** Visualization **Angella Kimuli Namyalo:** Investigation **Richard Muganzi:** Investigation **Patrick Ogwok:** Investigation **Claire Nankoma:** Investigation **Susan Nabadda:** Investigation **Tobias Alfvén:** Supervision **Juliet Mwanga-Amumpaire:** Supervision **Giulia Gaudenzi:** Conceptualization, Writing - Review & Editing, Funding acquisition, Supervision.

## Acknowledgments

Funding for this study was provided by the Swedish Research Council (Vetenskapsrådet, grant 2020-05396) and indirectly supported by the European Commission Horizon Europe Research and Innovation Program (Grant Agreement N°10105759). The authors acknowledge support from InfraVis for providing application expertise, data analysis and visualization resources for visualization through the Swedish Research Council grant 2021-00181.^23^ Some of the authors are affiliated with InfraVis, whose expertise in visualization infrastructure helped enable this collaboration. Generative AI tools were used for language and grammar editing.

## Data Sharing Statement

All collected data can be found at our OSF repository https://osf.io/j7puk/

